# Diversity in antibody titer at an average of 87 days after the second dose of BNT162b2 mRNA coronavirus disease 2019 vaccine: data from a predominantly elderly population in a practice in Tokyo

**DOI:** 10.1101/2021.12.22.21268296

**Authors:** Yasuko Fuse-Nagase, Mitsuo Nagase

## Abstract

It is very important for the elderly, who tend to have serious COVID-19 infection and high mortality rates, to maintain sufficient immunity. We reviewed the medical charts of predominantly elderly population to obtain the data on serum anti-SARS-CoV-2S antibody titer after complete vaccination with the BNT162b2 mRNA vaccine (two doses) and evaluated the background factors associated with the titer. We enrolled 230 participants (101 men and 129 women). Their average age was 71.9 (SD 12.5) years, and median was 72 years. The anti-SARS-CoV-2S antibody titer varied from 0.55 U/mL to 4920 U/mL. We found that the value of the titer varied widely. The value of the titer was negatively associated with age, alcohol consumption, time elapsed from second vaccine dose, and use of immunosuppressive medication. The result that the titer was negatively associated with aging suggests that the timing of additional shot should be carefully determined especially among elderly population.

## Introduction

Even though the BNT162b2 mRNA vaccine against COVID-19 has shown to be effective in controlling the COVID-19 pandemic in many countries[1][2], cases of breakthrough infection, i.e., COVID-19 infection despite complete vaccination have been reported[3]. Further, while the booster is also currently available in several countries, the optimal timing for its administration has not yet been clearly established. It is important for the elderly, who tend to have serious COVID-19 infection and high mortality rates[4], to maintain sufficient immunity.

The presence of the anti-SARS-CoV-2S antibody has been reported to be related to reduced risk of the infection[5]. However, previous studies on titer mainly targeted health-care workers and those who targeted elderly population is scares. We reviewed the medical charts of predominantly elderly population to obtain the data on serum anti-SARS-CoV-2S antibody titer after complete vaccination with the BNT162b2 mRNA vaccine (two doses) and evaluated the background factors associated with the titer.

## Methods

The participants of this study had no clinical history of COVID-19, had received both doses of the BNT162b2 mRNA vaccine in 2021, and were tested for serum anti-SARS-CoV-2S antibody titer at Nagase Clinic in Tokyo, Japan. All participant data were obtained from their medical charts including age, sex, history of drinking and smoking, time elapsed from the second vaccine dose to blood sampling, whether prescribed immunosuppressive medication, as well as the titer. The titer measurement was performed on clinical purpose to confirm they had sufficient immunity against SARS-CoV-2S between August–October 2021. It was outsourced to LSI Medience Corporation, a company that provides a clinical laboratory service, and they used the Elecsys Anti-SARS-CoV-2S RUO (Roche Diagnostics). Titer values ≥ 0.8 U/mL were defined as positive.

This study was approved by the ethics committee of Ibaraki University (N. 210300). Informed consent was obtained from all the participants in this study.

We used stepwise multivariate linear regression to identify factors associated with serum anti-SARS-CoV-2S antibody titer. Nominal data were converted to dummy variables and the titer value was used as the response variable. Explanatory variables were sex (female: 0, male: 1), age, time elapsed after second immunization (days), alcohol consumption (no: 0, occasionally: 1, almost every day: 2), smoking habit (non-smoke: 0, smoke: 1), and use of immunosuppressive medication (no: 0, yes: 1). Statistical Package for Social Sciences (SPSS, ver. 28) was used for statistical analyses; P < 0.05 was considered statistically significant.

## Results and discussion

We enrolled 230 participants (101 men and 129 women), and all participants were Japanese. Participant age ranged from 35 to 101 years, and 82.2% were 60 years or older. Average age was 71.9 ± 12.5 years (mean ± SD), while mode was 81 years, median was 72 years, and first and third quartiles (Q1–Q3) were 64 and 81 years, respectively. Next, while 18 participants consumed alcohol almost every day, 72 from time to time and 140 did not, 18 participants smoked and 10 required immunosuppressive medication for rheumatoid arthritis. The time from second dose of vaccine to blood sampling ranged from 27 to 137 days (mean, 86.7 ± 23.0 days; median, 87 days; Q1–Q3, 72 and 104 days, respectively).

The anti-SARS-CoV-2S antibody titer varied from 0.55 U/mL to 4920 U/mL and the distribution is depicted in Figure 1. The data showed a mode of 419.0 U/mL, median of 382.5 U/mL, and Q1–Q3 values of 197.8 U/mL and 789.3 U/mL, respectively. All but one of the participants had titer levels that were categorized as positive.

**Fig 1.** The distribution of the anti-SARS-CoV-2S antibody titer. The distribution of the anti-SARS-CoV-2S antibody titer varied widely from 0.55 U/mL to 4920 U/mL. The data showed a mode of 419.0 U/mL, median of 382.5 U/mL, and Q1–Q3 values of 197.8 U/mL and 789.3 U/mL, respectively.

The results of multivariate linear regression analysis are summarized in Table 1 and the independent factors associated with titer levels were age, alcohol consumption, time elapsed from second vaccine dose, and use of immunosuppressive medication.

**Table 1.** Summary of multivariate analysis to evaluate the factors associated with antibody titer.

| Variables | Standardized regression coefficient | 95% confidence interval |  | P value | VIF |
| --- | --- | --- | --- | --- | --- |
| Age | -0.28 | -19.89 | -6.88 | < 0.01 | 1.28 |
| Alcohol | -0.23 | -332.92 | -101.74 | < 0.01 | 1.05 |
| Days from the second shot | -0.19 | -8.46 | -1.42 | < 0.01 | 1.26 |
| Immunosuppressant | -0.15 | -797.42 | -88.6 | 0,02 | 1.01 |
| (Adjusted R2 = 0.19) |  |  |  |  |  |

We show that the anti-SARS-CoV-2S antibody titer varied widely even though all participants had received both vaccine doses and that age, longer time from second vaccine dose, alcohol consumption, and use of immunosuppressive medication were related to lower titer.

Median titer values reported by previous studies that used identical measurement methods are 2060.0 U/mL[6] and 764 U/mL[7], which are greater than our result of 419.0 U/mL. This difference may be attributed to older participants in our study compared to those in previous studies[6] [7] and to a comparatively longer duration between sampling and second vaccination dose[6].

The association between aging and lower titer is consistent with results from previous reports[6]–[9]. The association between alcohol consumption, time from the second vaccine dose, and use of immunosuppressive medication with lower titer are also consistent with the report by Kageyama et al[6].

One strength of our study is that we were able to analyze titer among the elderly, and we believe that our study would provide information useful for optimal scheduling of additional vaccinations. Our study also has a limitation. We excluded participants who had a clinical history of COVID-19, but our study cohort could have included some asymptomatic cases as we neither measured antibody titers before the first vaccination dose nor performed PCR to rule out infection.

## Conclusion

We found that the value of the titer varied widely. We also found that the value is negatively associated with some factors including aging and time from the second vaccine dose. As the elderly tend to have serious COVID-19 infection and high mortality rates^4^, the timing of additional shot should be carefully determined.

## Data Availability

All data produced in the present study are available upon reasonable request to the authors.

## Acknowledgement

We acknowledge the staff of Nagase Clinic for their co-operation.

